# A Susceptible Vaccinated Exposed Infected Hospitalized and Removed/Recovered (SVEIHR) Model Framework for COVID-19

**DOI:** 10.1101/2023.08.10.23293942

**Authors:** S.O. Oyamakin, J.I Popoola

## Abstract

In reaction to the severe socio-economic effects and upheavals that the Covid-19 sickness had on the world within the first few weeks of its introduction, everyone involved had to act quickly to look for possible solutions for preventing the ensuing epidemics. A prompt response is more critical given Nigeria’s subpar social, economic, and healthcare infrastructure. Investigated was the efficacy of various pharmacological, non-pharmaceutical, or a combination of both therapies in flattening the Covid-19 incidence curve. In order to investigate the impact of these interventions, a deterministic SVEIHR model was created and applied. The Nigerian Center for Disease Control (NCDC) portal’s Covid-19 data were used to parametrize the model. For simulations using a system dynamic simulation, estimated parameters were employed. The fundamental reproduction number, R_0_, was used to evaluate the success of our suggested intervention in effectively managing COVID-19 transmission. The simulation results demonstrated that the use of only non-pharmaceutical interventions, such as the use of face masks, a light lockdown, and hand washing at baseline or high levels, is insufficient, with the R_0_ varying from vaccination at the vaccination rate of 0.5% with non-pharmaceutical interventions at any level of compliance, and a combination of vaccination at 0.05% and high hygiene level were effective in flattening the Covid-19 disease incidence curve in Nigeria, returning a R_0_ less than 0. Furthermore, maintaining a high level of cleanliness, which includes hand washing and the use of a face mask, would be sufficient to stop the spread of Covid-19 disease and eventually flatten Covid-19 disease incidence curve in Nigeria, given a low turnout of 0.05% for vaccination and the easing of lockdown.

## Introduction

The world was somewhat better prepared for covid-19, a zoonotic infectious disease caused by the severe acute respiratory syndrome corona virus, when it first appeared in Wuhan, China in the middle of December 2019 and later spread throughout the world as both pharmaceutical and non-pharmaceutical interventions were swiftly implemented. In record time, vaccinations were also produced and tested. As of April 4, 2023 (WHO Coronavirus Dashboard; www.covid19.who.int), there have been 762,201,169 confirmed cases and 6,893,190 fatalities worldwide since the World Health Organization declared it a pandemic on March 11, 2020 (Ngonghala et al., 2020; World Health Organization, 2020a; World Health Organization). Nigeria first discovered its index case on February 25, 2020, in Lagos. Following that, a contact of the index was discovered in Ewekoro, Ogun State, Nigeria, and it quickly spread to all of the states in the nation.Few states, such as Kano, hesitated to admit the sickness existed there until certain tragic deaths that may have been caused by the ailment were documented.

Prior to doses of the popular vaccines from AtsraZeneca and Johnson & Johnson being given in Nigeria, various non-pharmaceutical intervention policies with varying degrees of enforcement and adherence were put in place. Each had a different effect on the progression of the disease with varying degrees of effectiveness. Estimating the incubation period—the time between catching the illness and exhibiting symptoms—early enough will help determine the number of days required for isolation. The longest incubation duration, which has since been adopted, was calculated by Jiang et al. (Jiang et al, 2019) to be 14 days.

In a review of 12 research, Liu et al. (Liu et al., 2020) found that the basic reproduction number for the covid-19 virus originated in China and a small number of other countries that underwent early diffusion. They obtained estimates ranging from 1.4 to 6.49, with a mean of 3.28 and an interquartile range of 1.16.

To better understand the dynamics of the covid-19 disease’s transmission, several models have been created. Both stochastic and mathematical models are included in these models. To assess the impact of public health initiatives on the dynamics of disease transmission while taking into account under-reporting in the Ivory Coast, Coulibaly and N’zi (2020) created a stochastic model with jump hinged on the deterministic SIRU where U stands for unreported. The use of face masks in conjunction with other non-pharmaceutical methods is beneficial in reducing disease burden and preventing community transmission, according to Eikenberry et al.’s assessment of the impact of mask use by the asymptomatic population in China in 2020. Using a baseline model with the Susceptible S(t), Exposed E(t), Asymptomatically infected A(t), Symptomatically infected, Quarantined Q(t), Hospitalized H(t), and Recovered R(t) compartments, Ngonghala et al. (Ngonghala et al., 2020) also suggested that the use of surgical facemasks with efficacy greater than 70%. Adewole et al. (Adewole et al., 2021) examined the efficacy of three different tactics on the eradication of the infected population in Nigeria in order to predict the dynamics of COVID-19 transmission in Nigeria using an SEIR model with an additional compartment for quarantined. They concluded that each method alone is insufficient for containing the disease but is more effective when used in tandem with other strategies, with the appropriate stakeholders being required to assure compliance. Additionally, Daniel (Daniel, 2021) suggested an SEIR model (with additional compartments for the quarantined and critically ill population) that included both direct and indirect transmission paths to study the effects of a few non-pharmaceutical treatments on the disease transmission in Nigeria. Iboi et al. (Iboi et al., 2020), in their work on mathematical modeling and analysis of COVID-19 in Nigeria, proposed a mathematical model that estimated the fundamental reproduction number R0 as 2.13 and then investigated measures like various levels of social distance and public face mask wearing that could eventually make R0 1. System dynamics were employed by Jiang et al. (Jiang et al., 2022) to evaluate the efficacy of societal mobilization and immunization programs for Covid-19 in the United States. Using a modified SEIR model with additional compartments for those who have received vaccinations and those who have been hospitalized, this study examined the impact and effectiveness of pharmaceutical and non-pharmaceutical measures, or a combination of both, in flattening the covid-19 disease incidence curve.

### Model formation

The SEIR model used in this study has been modified to account for the portion of the population that has received vaccinations and those who are hospitalized, creating an SVEIHR model. particular as N(t), the total population at a particular moment t is partitioned into non-overlapping compartments. The susceptible population is represented by (S), and the susceptible individuals who have been immunized (have received at least one dose of the vaccine) enter the compartment of the immunized persons (V). They become exposed (E) when susceptible and immunized individuals have meaningful contact with those who are already sick. At the end of the incubation period, they may then either be infectious with symptoms (*I*_*s*_) or infectious without symptoms of illness (*I*_*a*_). While gravely ill and exhibiting symptoms, infectious individuals can eventually recover (R).

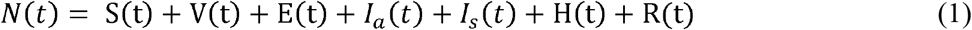

#### Model equations

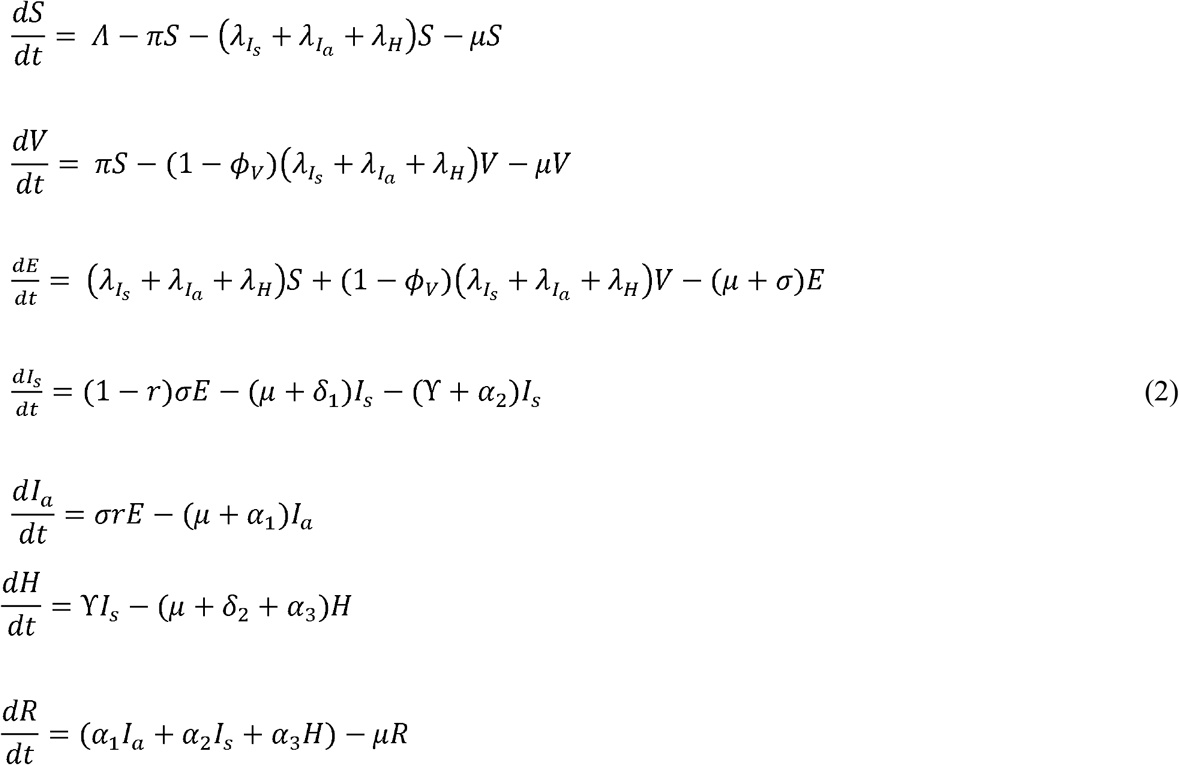

Where;

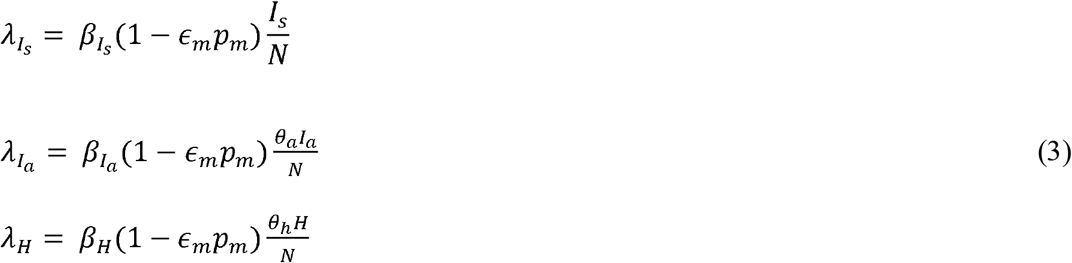

Following a successful interaction with an infected person in compartments *I*_*a*,_*I*_*s*,_*H* at rates 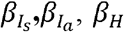, vulnerable persons who have not been immunized and few immunized individuals get exposed. Following the incubation period, exposed persons become contagious whether they exhibit symptoms or not. Only infectious patients who exhibit symptoms are eligible for hospitalization. Only those who were symptomatically contagious and, in a hospital, may finally pass away from the illness.

**Table 1:** Description of Model Parameters.

| Parameter | Description |
| --- | --- |
| $\Lambda$ | Recruitment rate into the susceptible compartment. Recruitment is by birth or migration |
| $\pi$ | Vaccination rate. |
| $\mu$ | Death rate unconnected to COVID-19 |
| $\delta_1, \delta_2$ | Death rate connected to COVID-19 |
| $\alpha_1, \alpha_2, \alpha_3$ | Recovery rates from $I_a$, $I_s$ and H compartments respectively |
| $\Upsilon$ | Hospitalization rate for infectious individuals |
| $\phi_v$ | Vaccine efficacy |
| $r$ | Proportion of the exposed, who are asymptomatic at the end of the incubation period |
| $(1 - r)$ | Proportion of the exposed, who show symptoms of the infection at the end of the incubation period |
| $\sigma r$ | Rate of progression from exposed to asymptomatic compartments |
| $(1 - r)\sigma$ | Rate of progression from exposed to infectious compartments |
| $\beta_{I_s}, \beta_{I_a}, \beta_H$ | Effective contact rate between those susceptible and the population in $I_s$, $I_a$ and H compartments respectively |
| $p_m$ | Proportion of the population wearing mask |
| $\epsilon_m$ | Efficiency of face mask in preventing acquisition of infection |
| □ | Proportion of the population that wash their hands |
| $\lambda_{I_s}, \lambda_{I_a}, \lambda_H$ | Forces of infection |
| $\theta_a, \theta_h$ | Reduced transmission factor for the asymptomatic and hospitalized. |

#### Properties of the model

##### Positivity of solutions

This is to show that given the initial conditions S(0)>0, V(0)>0, E(0)>0, *I*_*s*_ (0)> 0, *I*_*a*_ (0)> 0, H(0)>0, R(0)>0, the solution set S(t), V(t), E(t), *I*_*s*_(t), *I*_*a*_(t), H(t), R(t) are non-negative for all t>0.

Recall that from (3.19)

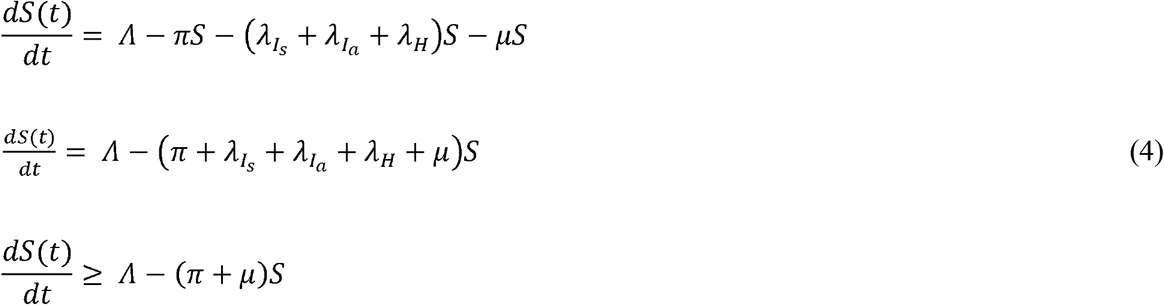

Using integrating factor;

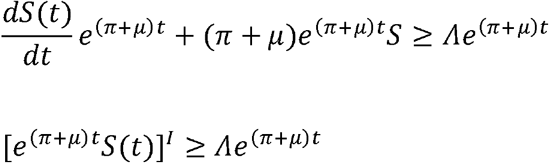

Integrate both sides;

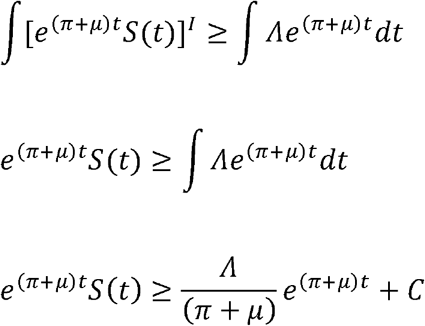

To find C, set t=0;

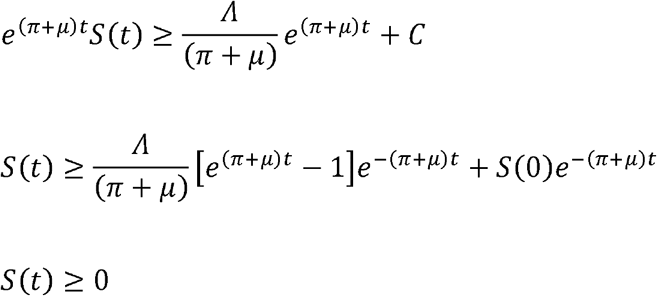

Additionally, it can be demonstrated that, given the initial values, all other state variables are positive for any t > 0.

##### Biological Equilibrium Point for Covid-19

In the Covid-19 disease-free state, the illness is absent, hence all afflicted classes (E, Is, Ia, and H) have a value of zero. We zero out the system of equations in (2) in order to get expressions for all states at the disease-free equilibrium point.

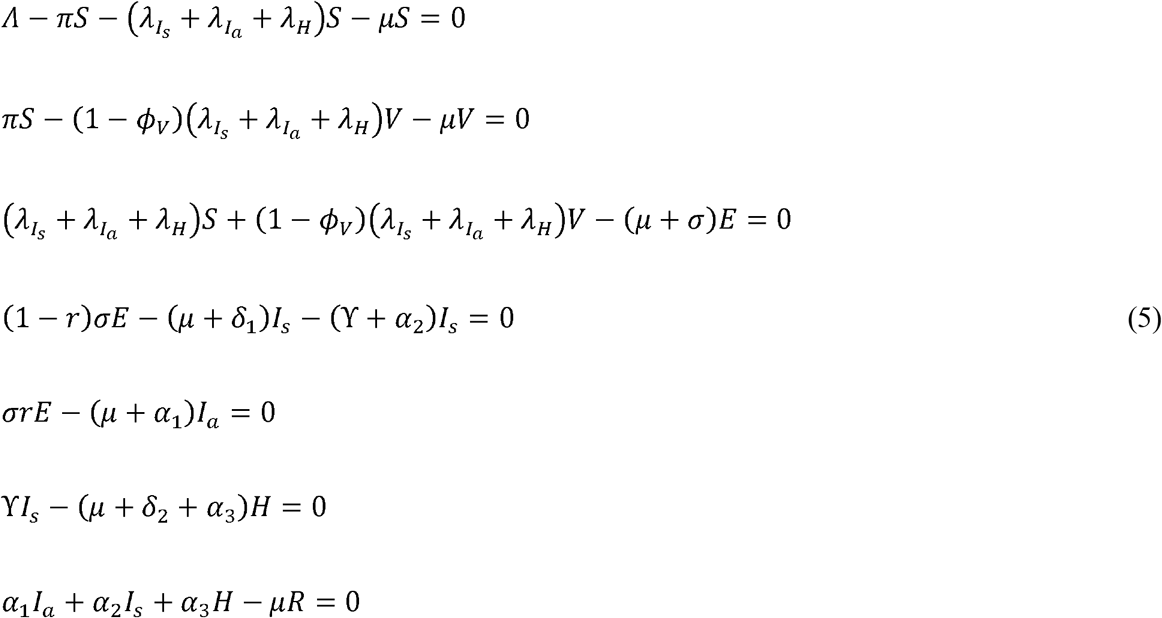

Substituting *E, I*_*s*_, *I*_*a*_, *H* = 0 in the system of equations (4), the disease-free equilibrium point denoted as;

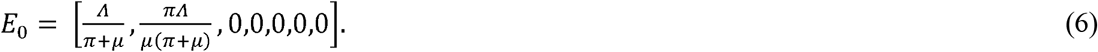

##### Stability of the disease-free equilibrium point

The disease-free equilibrium point is asymptotically stable if the Jacobian of (3.1) is obtained at the disease-free equilibrium point, 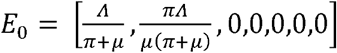 and 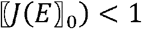. The Jacobian matrix was deduced using the maple software as given below;

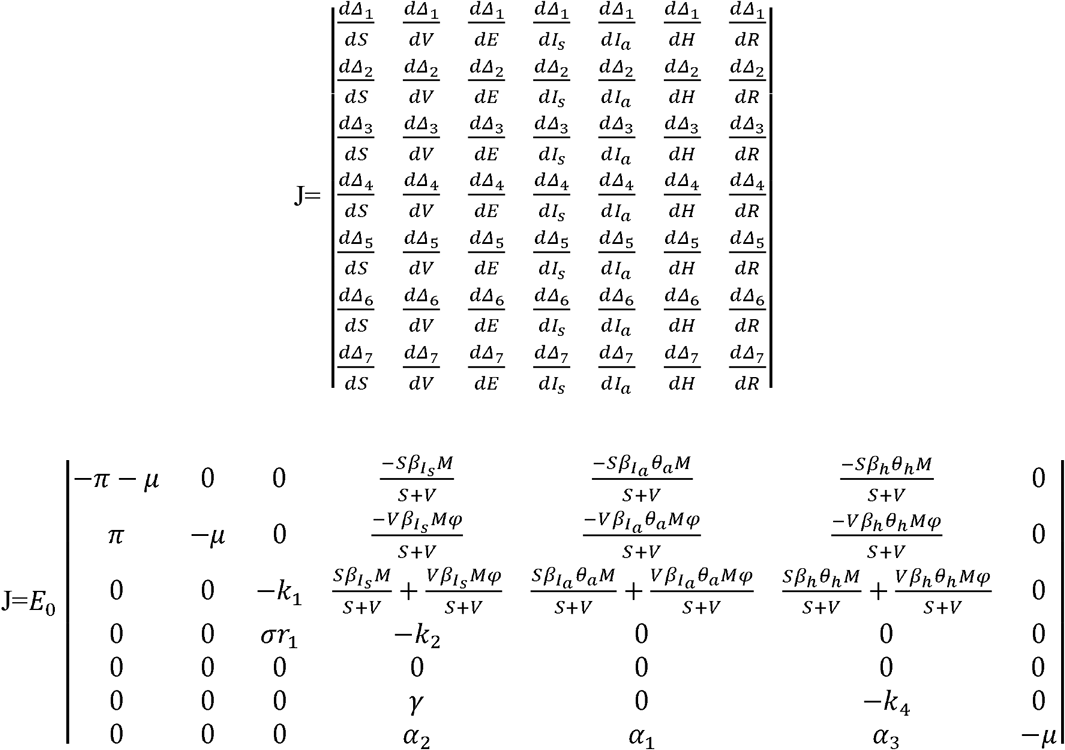

Where;

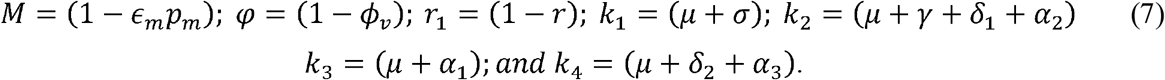

Next step was that we established the eigenvalues of the Jacobian at the disease-free equilibrium point, by obtaining |*J*(*E*_0_) − *λI*|, where I is the 7×7 identity matrix. The second and seventh columns contain only the diagonal entry which gave one negative eigenvalue, -μ. Others can be found by deleting the second and seventh rows and columns.

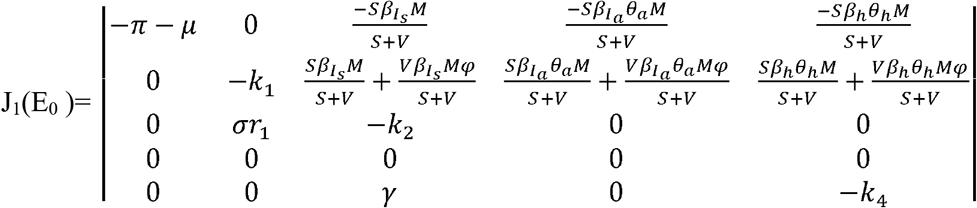

The first column contained only the diagonal entry which gave another eigenvalue (-π-μ). We proceeded by deleting the first column and first row in J_1_(E_0_) to obtain other eigenvalues.

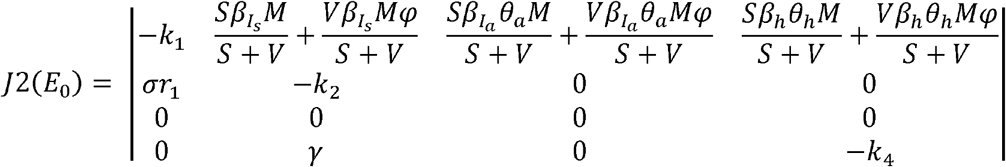

Upon solving, the eigenvalues were found to be negative which implied that the disease-free equilibrium point is locally stable.

#### Endemic Equilibrium Point

The steady-state solution in areas with disease is known as the endemic equilibrium points. The system of equations (4) and denote the endemic equilibrium points as follows are used to derive the endemic equilibrium points;

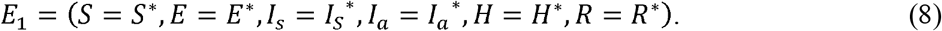

Using maple software, the following result was gotten;

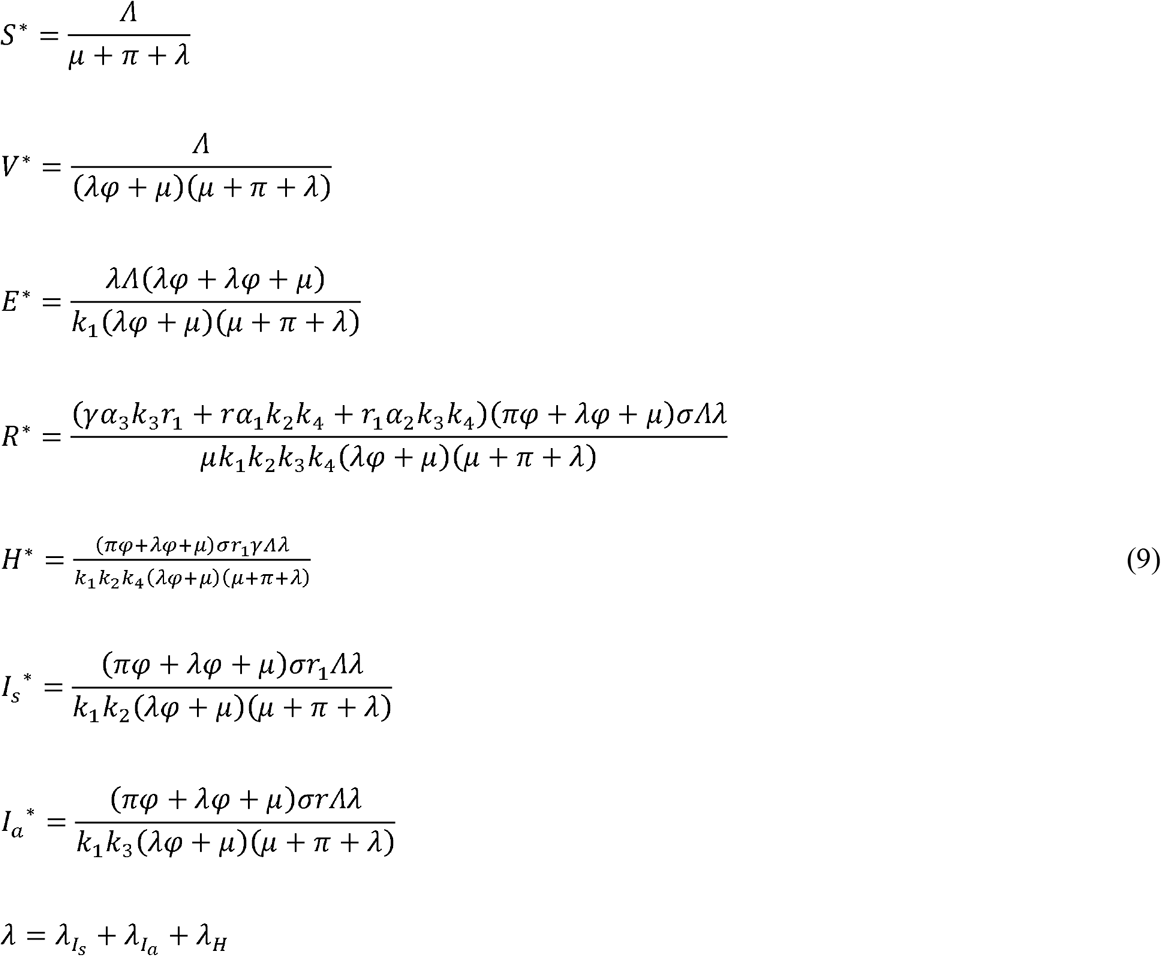

#### Basic Reproduction Number

To prove the local stability of the disease-free equilibrium point, it was necessary to derive an expression for the basic reproduction number which was done using the next generation matrix method. The Jacobian matrices F and V, representing the new infection terms and the transfer terms was deduced. The infective states are. *E, I*_*s*_, *I*_*a*_, *H*.

For F, Jacobian of the following system of equations below is obtained.

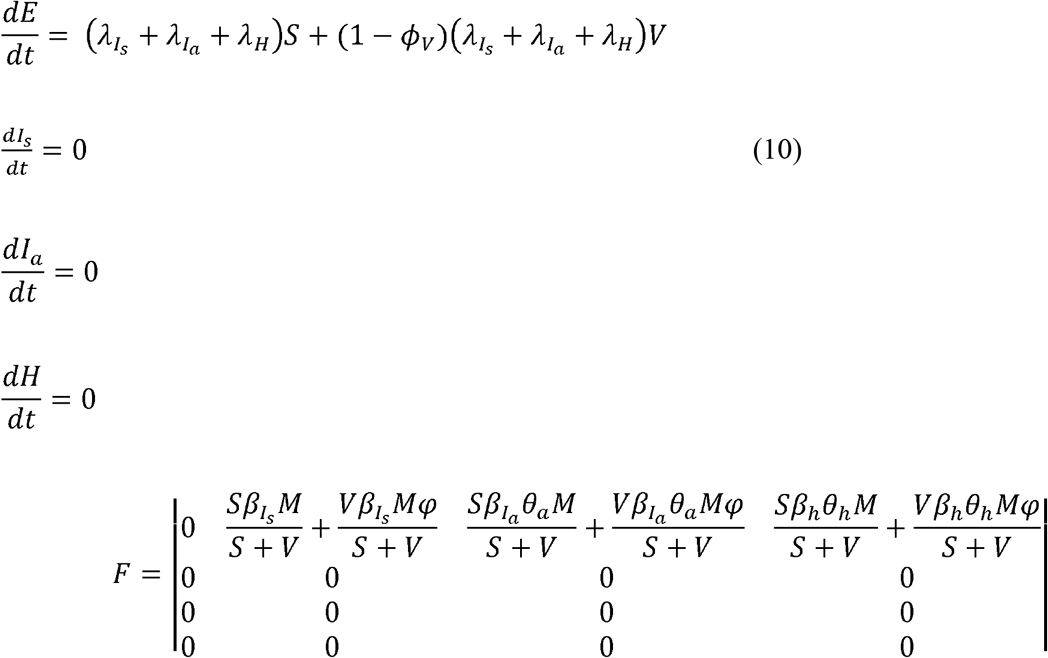

For V, the Jacobian of the system of equations (showing the transmission state) which is given below is obtained.

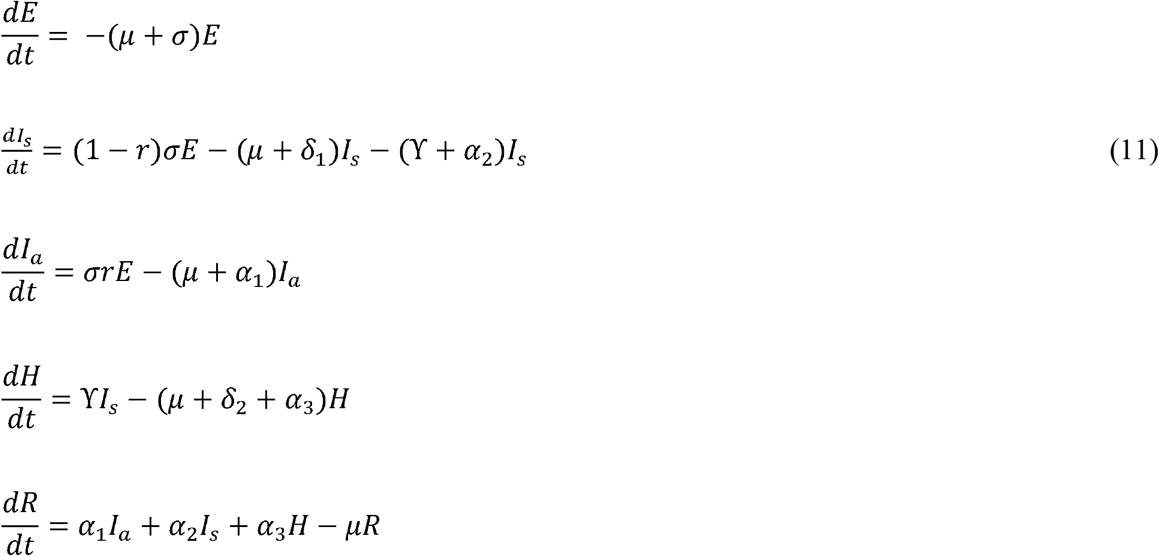

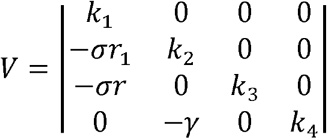

By resolving the spectrum radius (dominant eigenvalue) of *FV*^−1^, the fundamental reproduction number R_0_ (average number of secondary infections produced by an infectious individual in a fully susceptible population during the entire period of infectiousness) is obtained.

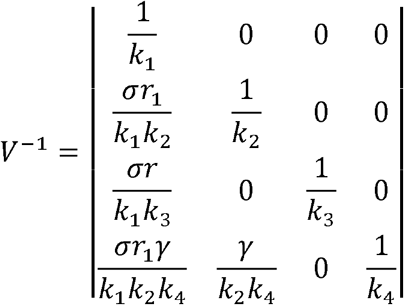

After evaluating the spectral radius of *FV*^−1^ four eigenvalues were obtained as follows;

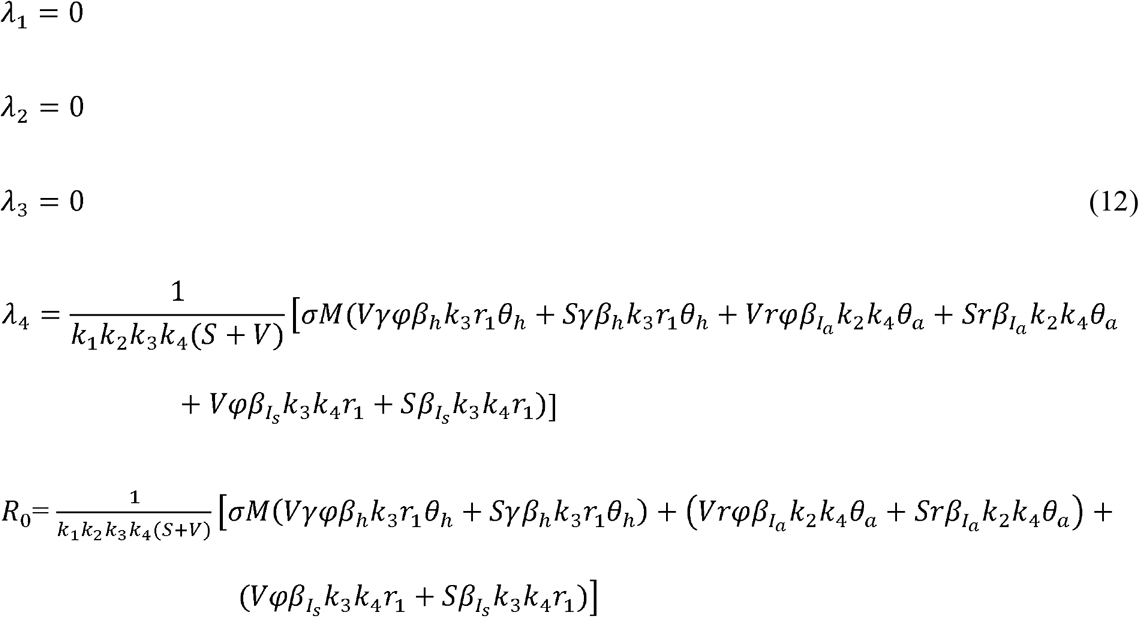

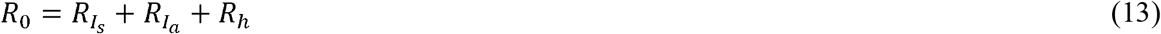

##### Numerical result

Using data from the Covid-19 reported incidence in Nigeria as reported by the NCDC from February 28, 2020, to March 1, 2021, before immunization, parameters were computed using Mathematica software. A system of nonlinear differential equations can be solved using Mathematica’s ParametricNDSolve function, which also provides an estimate for the parameters that are inputted as one of the function’s arguments.

##### Parameter values and data fitting

A few of the model’s initial parameters were sourced from the literature, while others were predicated on well acknowledged facts concerning Covid-19 in Nigeria (Iboi et al., 2020; Deborah 2020; Adewole et al., 2021). Using data on Covid-19 incidence in Nigeria from 28 February 2020 to 1 March 2021 prior to immunization (Nigeria Centre for Disease Control, 2021; World Health Organization, 2021), nine factors were estimated.

**Table 2:**
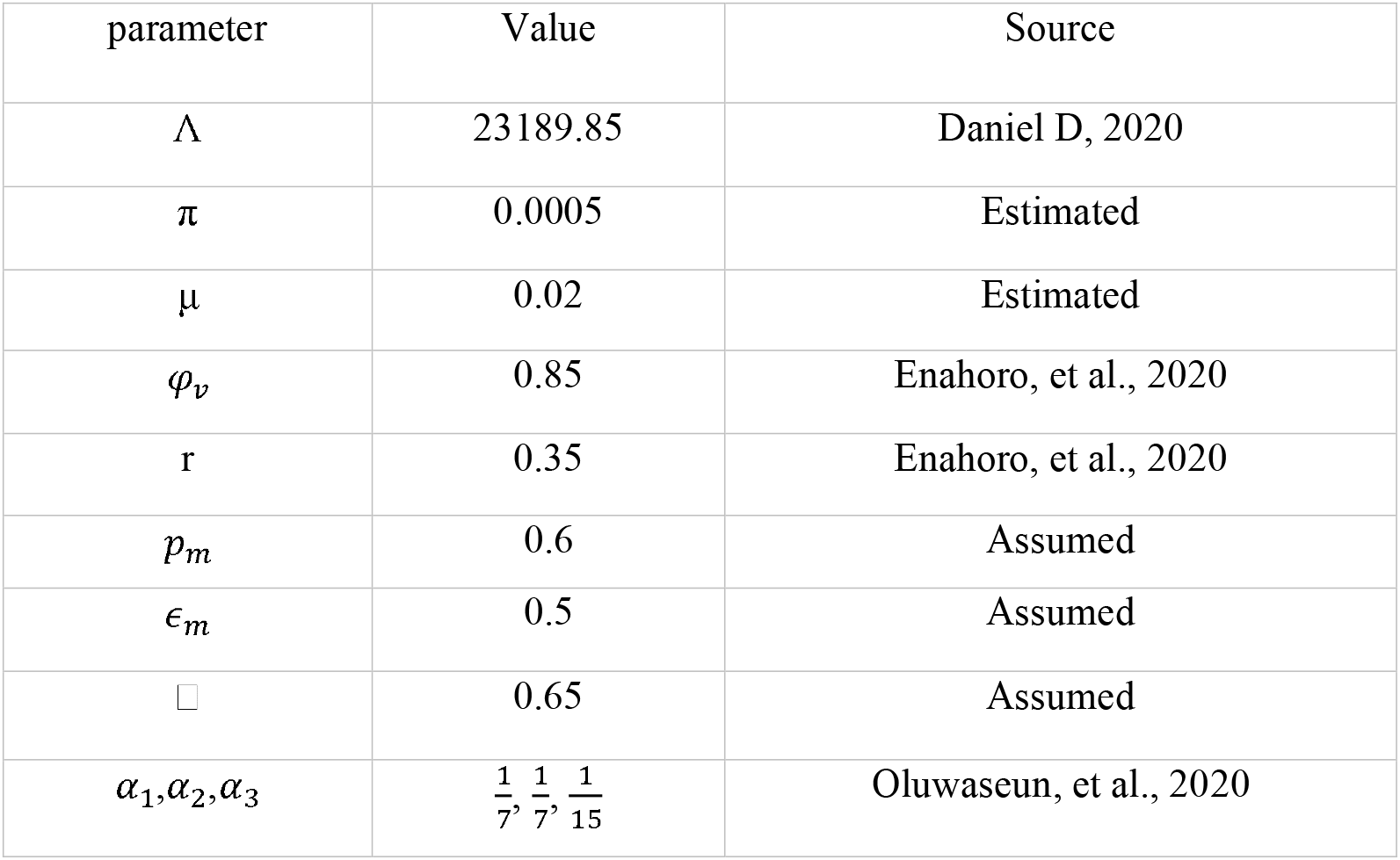
Parameter values taken from literature/estimated.

**Table 3:** Parameter values from data fitting with 95% confidence interval.

| Parameter | Value | C.I |
| --- | --- | --- |
| $\delta_1$ | 0.288 | [0.243, 0.291] |
| $\delta_2$ | 0.151 | [0.141, 0.238] |
| $\sigma$ | 0.125 | [0.12, 0.133] |
| $\beta_s$ | 1.082 | [0.825, 1.1] |
| $\beta_a$ | 0.302 | [0.288, 0.344] |
| $\beta_h$ | 0.108 | [0.937, 0.116] |
| $\theta_a$ | 0.74 | [0.535, 0.854] |
| $\theta_h$ | 0.45 | [0.332, 0.511] |
| $\gamma$ | 0.154 | [0.069, 0.183] |

The basic reproduction number R_0_ was determined after the execution of each preventative measure, which might be non-pharmaceutical, pharmacological in the form of vaccination, or a combination of both kinds of measures, to determine the impact of the measure in slowing the Covid-19 outbreak.

## Results of simulation

Using Vensim, a program for simulating and deploying models of complex dynamic systems, the model 2.1 is examined to determine the population-level impact of each suggested countermeasure in flattening the Covid-19 illness incidence curve in Nigeria. Proposed interventions are those that are thought to be implementable in light of Nigeria’s socioeconomic conditions. Step size is 380 days.

**Figure 1:**
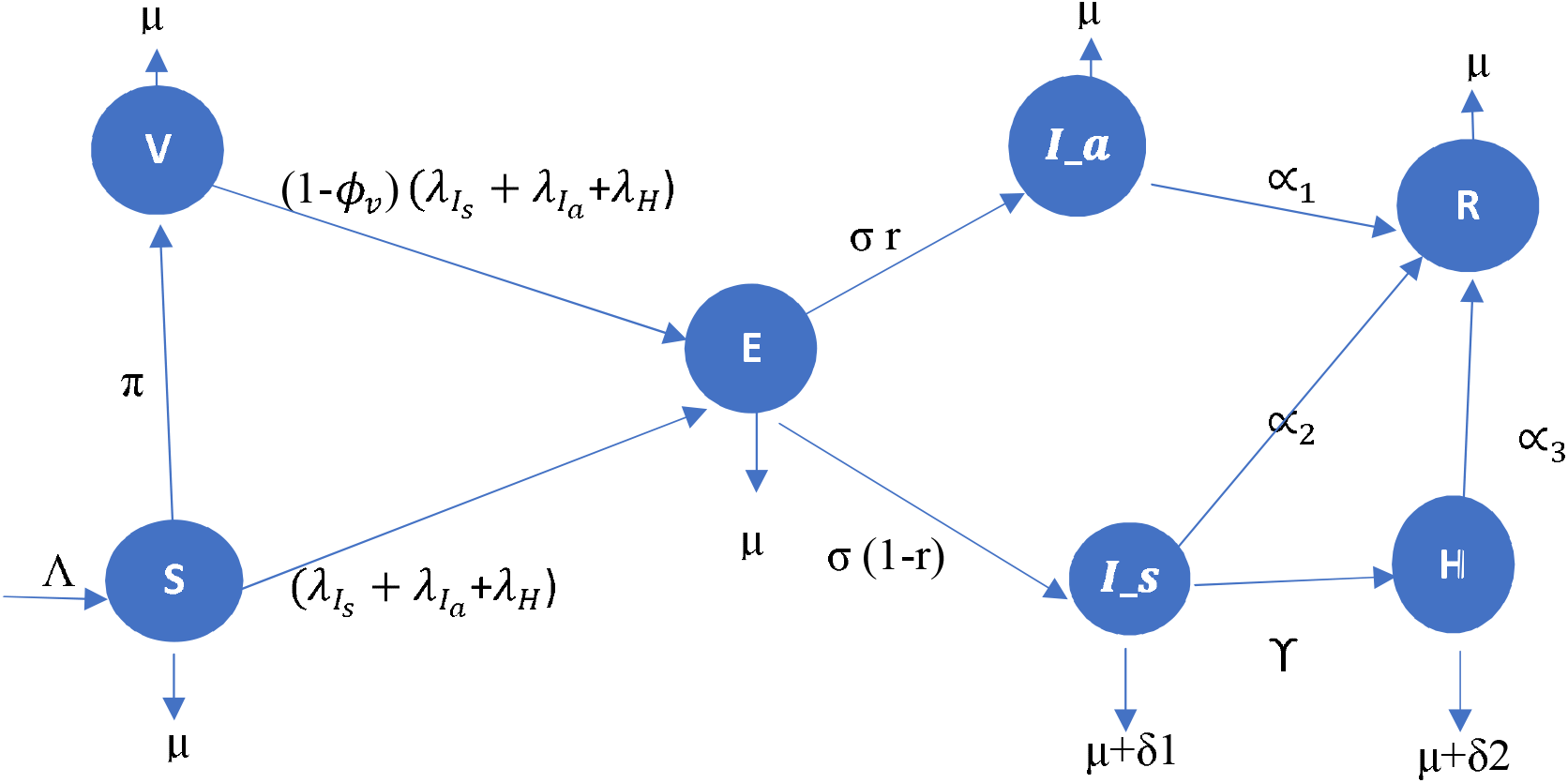
Schematic diagram of the proposed SVEIHR model.

**Figure 2:**
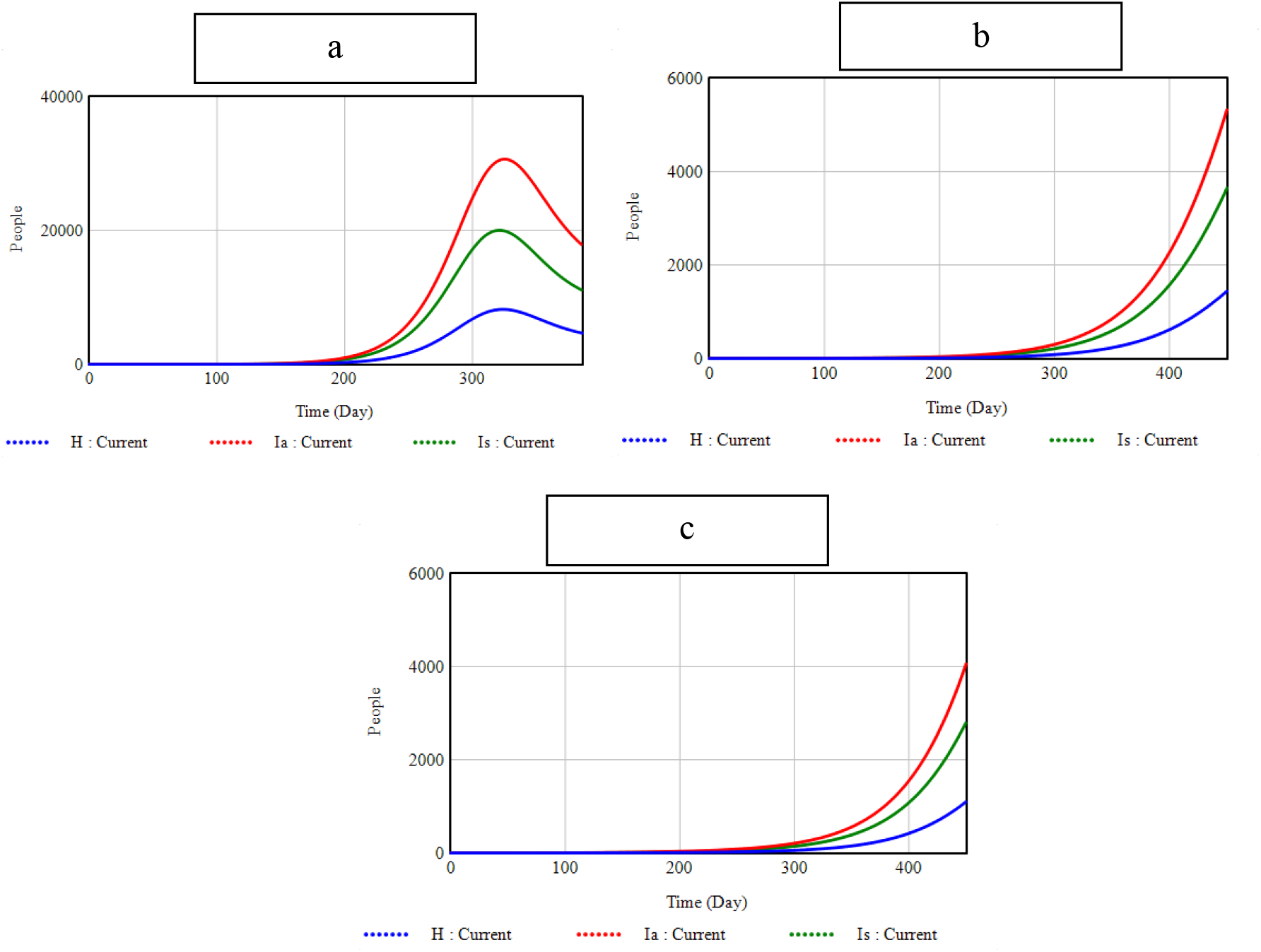
(a) Simulation showing incidence with parameters at baseline level; (b) washing of hands and wearing of face mask, *P*_*m*_ = 0.75 with an effectiveness of *ε*_*m*_= 0.65 while = 0.8. (c) Mild social distancing, effective contact rates *β*_*i*_, *β*_*a*_, *β*_*h*_ were reduce by 10% while all other parameter values are at baseline level.

### Non-pharmaceutical interventions

Non-pharmaceutical treatments may be able to reduce disease incidence, but Covid-19 disease incidence after their adoption is still very high and the epidemic has not been stopped, as can be shown from R_0 values. Social separation appears to be the non-pharmaceutical strategy that is most significantly reducing the occurrence of the Covid-19 sickness in Nigeria.

**Figure 3.**
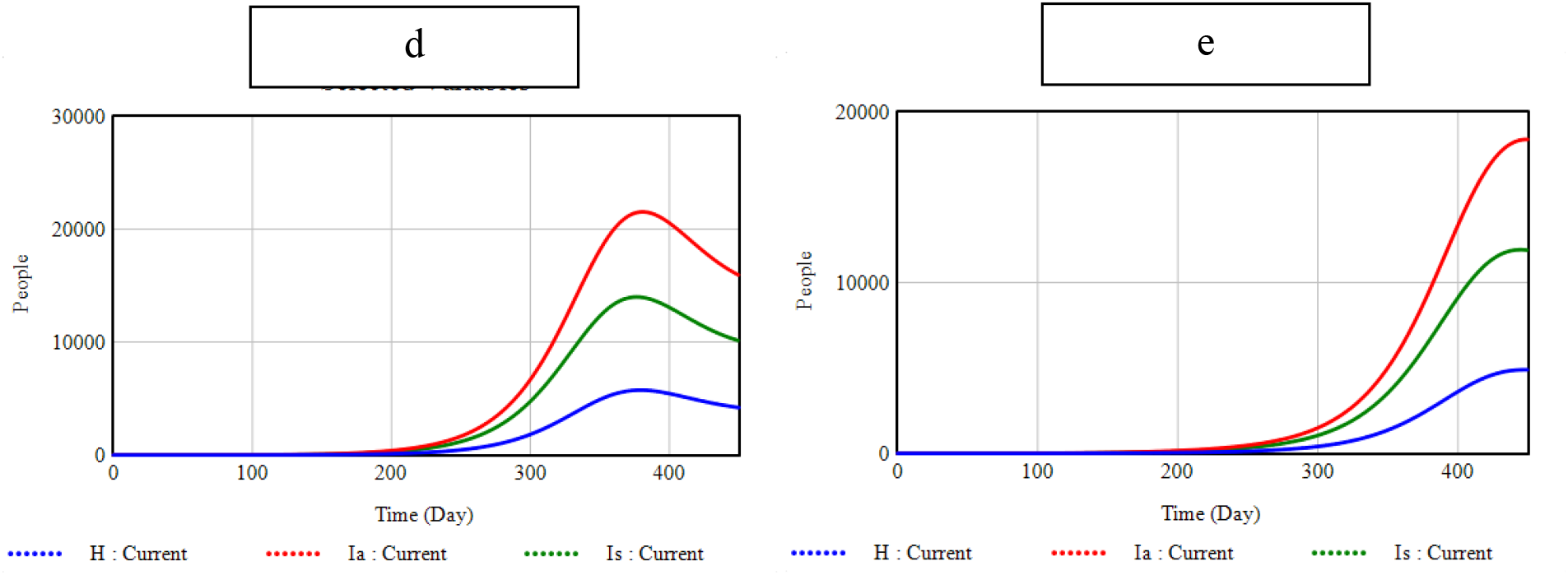
**(d) Vaccination, where vaccination rate π = 0. 0005, vaccine efficsyc *φ***_***ν***_ **= 0.85 and all other parameters kept at baseline level. (e) Increased vaccination rate, *π* is increased to 0.005.**.

**Figure 4.**
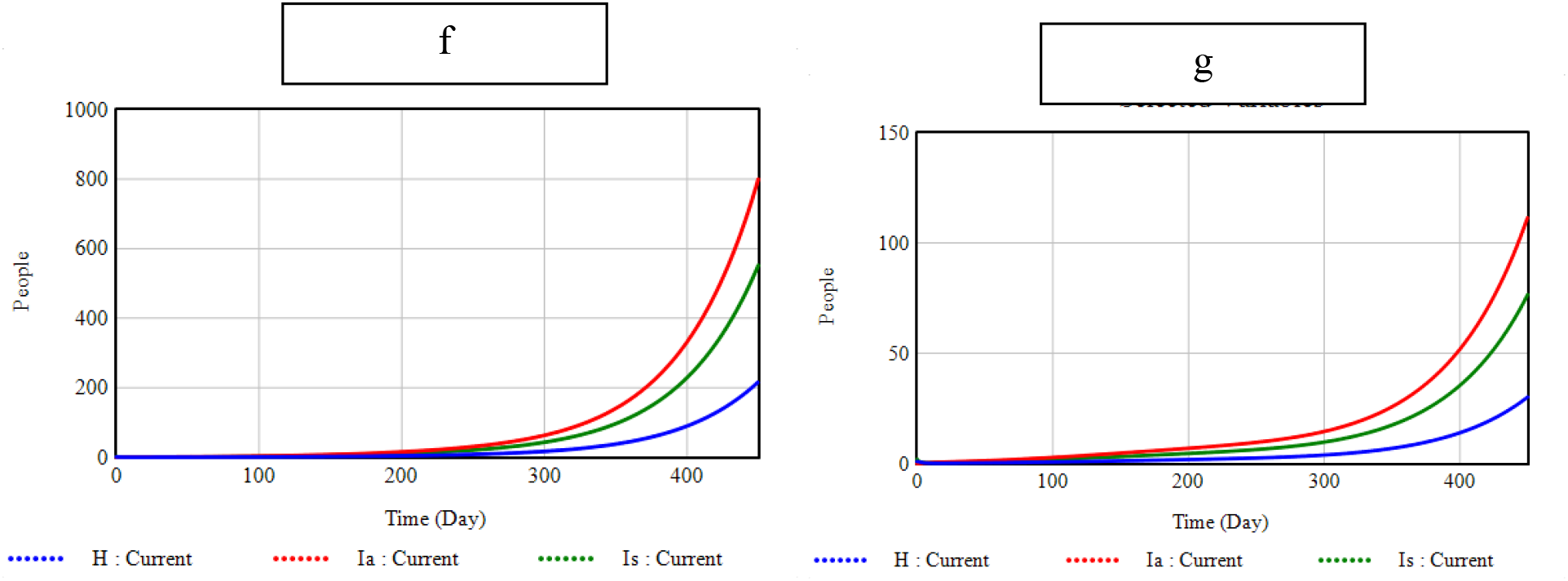
(f) High-level hygiene and vaccination, *P*_*m*_ = 0.75 with an effectiveness of *ε*_*m*_= 0.65 while = 0.8, vaccination rate π = 0. 0005 vaccine efficsyc *φ*_*ν*_ = 0.85. (g) Increased vaccination rate and high-level hygiene, *P*_*m*_ = 0.75 with an effectiveness of *ε*_*m*_= 0.65 while = 0.8, vaccination rate π = 0. 005, vaccine efficsyc *φ*_*ν*_ = 0.85

### Pharmaceutical intervention alone

Considered vaccination rates may reflect the attitudes of many Nigerians against immunization, which may discourage many from accepting to receive immunizations.

### Combination of pharmaceutical and non-pharmaceutical intervention

The table 4 below showed the percentage reduction in maximum number of persons that could be hospitalized in a single day.

**Table 4:**
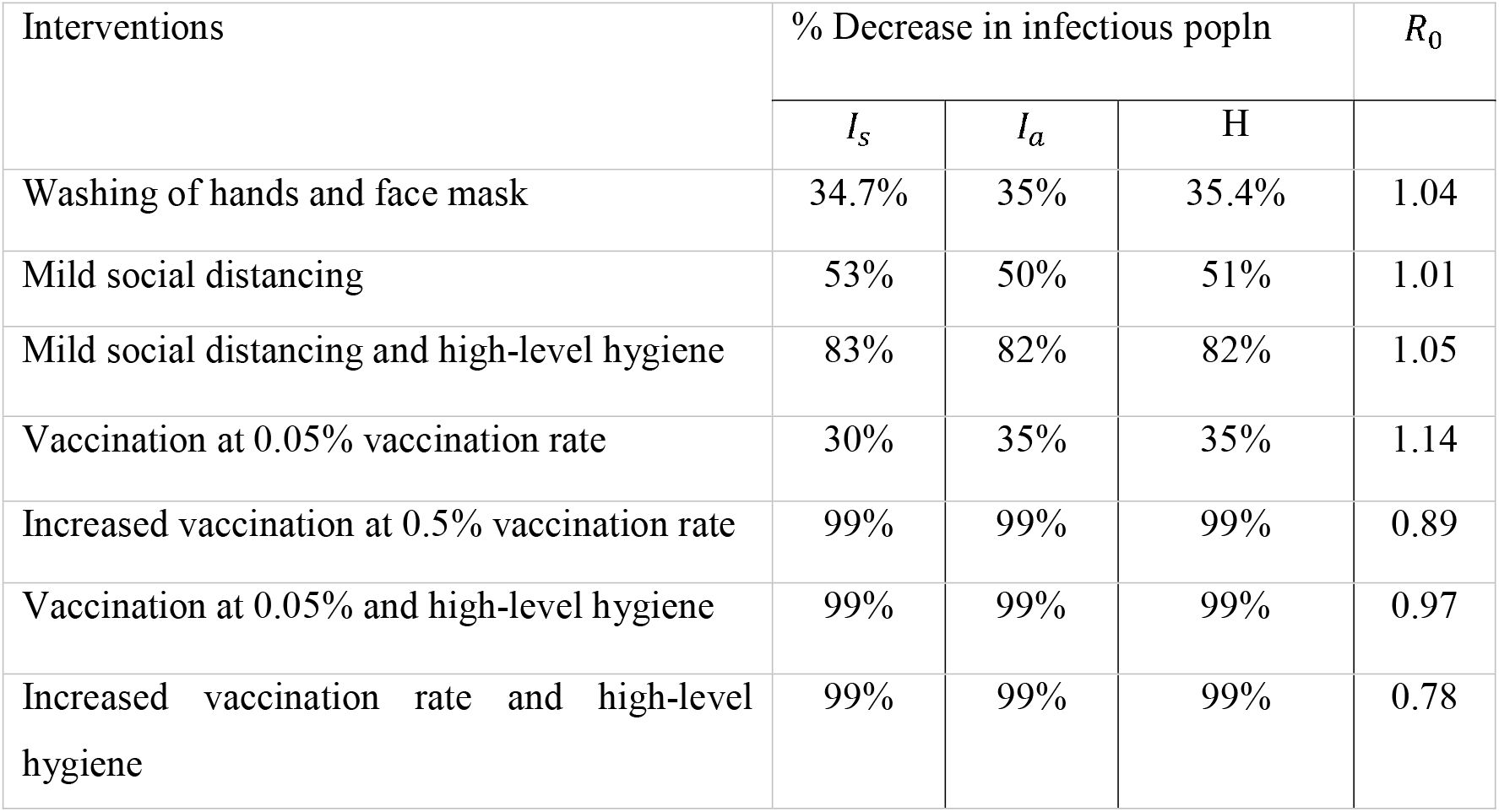
Percentage decrease in maximum incidence and *R*_0_.

| Interventions | % Decrease in infectious popln | | | $R_0$ |
| --- | --- | --- | --- | --- |
| | $I_s$ | $I_a$ | H | |
| Washing of hands and face mask | 34.7% | 35% | 35.4% | 1.04 |
| Mild social distancing | 53% | 50% | 51% | 1.01 |
| Mild social distancing and high-level hygiene | 83% | 82% | 82% | 1.05 |
| Vaccination at 0.05% vaccination rate | 30% | 35% | 35% | 1.14 |
| Increased vaccination at 0.5% vaccination rate | 99% | 99% | 99% | 0.89 |
| Vaccination at 0.05% and high-level hygiene | 99% | 99% | 99% | 0.97 |
| Increased vaccination rate and high-level hygiene | 99% | 99% | 99% | 0.78 |

## Conclusion

We won’t soon forget the effects that the Covid-19 sickness had on the entire planet. It certainly had a significant impact on how epidemics should be managed and taught the world how to react to future outbreaks of infectious diseases, from its effects on the global economy and social-cultural life of people to the intense shock it caused the public health sector of the very developed countries and the mortality experienced worldwide. Using a proposed SVEIHR compartmental model and simulation methodology carried out in a system dynamic simulation tool, this study examined the impact of non-pharmaceutical, pharmaceutical, or a combination of both measures in flattening the curve of Covid-19 disease in Nigeria.

Deterministic disease models have a few theoretical underpinnings, including as the solutions’ positivity, the invariant region, the equilibrium points, and justifications for their local stability.

The Nigerian Centre for Disease Control (NCDC) portal’s Covid-19 data on daily new cases was used to estimate model parameters under the assumption that the data was accurate.

After each applied intervention, the estimated basic reproduction number shown in Table 4 demonstrates that non-pharmaceutical measures at the recommended level of adherence are insufficient to halt the epidemic when used individually. Additionally, daily vaccination of 0.05% of the population is not a successful intervention strategy. To stop the epidemic and flatten the disease incidence curve, vaccination at a vaccination rate of 0.05 and combinations of both vaccination at any of the investigated vaccination rates are successful.

The simulation findings also indicate a noticeable decrease in the number of daily new cases that might be noted when the intervention measures are put into place. Therefore, it could be argued that a major decline in Covid-19 incidence does not necessarily herald the end of the Covid-19 epidemic in Nigeria.

## Data Availability

Some parameter estimates were obtained from literature and properly referenced. others were estimated by our proposed model

## Notes

### Competing Interest Statement

The authors have declared no competing interest.

### Funding Statement

None

